# The Impact of Midodrine On Guideline-Directed Medical Therapy in Patients Admitted with Systolic Heart Failure

**DOI:** 10.1101/2023.04.21.23288945

**Authors:** Christopher B Scoma, Dae Hyun Lee, David Money, Gerry Eichelberger, Ahsan Usmani, Adam J Cohen, Joel Fernandez

## Abstract

**Background:** Midodrine is occasionally used off-label to treat hypotension associated with advanced heart failure. Its association with changes in prescription of guideline-directed medical therapy (GDMT) has not previously been evaluated.

**Objectives:** We sought to evaluate the clinical characteristics and GDMT prescriptions of heart failure patients who were prescribed midodrine.

**Methods:** We performed a retrospective cohort study identifying all patients admitted to our hospital in 2020 with decompensated systolic heart failure who were prescribed midodrine upon discharge. They were compared to decompensated systolic heart failure patients who were not prescribed midodrine. Baseline characteristics, GDMT adjustments, and clinical outcomes were collected.

**Results:** 114 patients met inclusion criteria in the midodrine group and were compared to 358 patients in the control group. At baseline, the midodrine group had worse left ventricular function, more right ventricular dysfunction, and more severe heart failure symptoms. At 6-months, the midodrine group had more initiation or up-titration of beta blockers (24.6% vs.15.4%; p=0.035), renin-angiotensin-aldosterone system (RAAS) inhibitors (34.2% vs. 24.0%; p=0.043) and sodium-glucose cotransporter-2 inhibitors (SGLT2i) (19.3% vs.10.6%; p=0.024) compared to the non-midodrine group, with a similar pattern for MRA (mineralocorticoid receptor antagonists) prescriptions (17.5% vs. 11.5%; p=0.126). Mortality was not statistically different between the two groups, but the midodrine group had more frequent re-hospitalization for heart failure (39.5% vs. 25.4%; p=0.006).

**Conclusions:** Midodrine is frequently prescribed to patients presenting with systolic heart failure; the patients given midodrine tended to have more advanced heart failure and worse 6-month clinical outcomes. However, the patients who were prescribed midodrine achieved better initiation and up-titration of GDMT at 6 months compared to those who were not prescribed midodrine. Future prospective clinical trials are warranted to confirm these findings and determine if this translates to improved clinical outcomes.

## Introduction

Congestive heart failure (CHF) is a common cause of morbidity and mortality in the United States, with an estimated prevalence of over 5 million patients and causing more than one million hospitalizations per year.^1^ The prevalence has continued to increase over time, in part owing to an aging population; thus, in patients with systolic dysfunction, it is critical to initiate guideline-directed medical therapy (GDMT) which has been proven to prolong life and reduce re-hospitalizations.^2,3^ National GDMT utilization remains poor, with the majority of heart failure patients not receiving contemporary quadruple-therapy and fewer still on target doses.^4^ One of the most common limiting factors to GDMT initiation and titration is hypotension (both at baseline and medication-induced), particularly with beta blockers and angiotensin receptor-neprilysin inhibitors (ARNIs).^5,6^ Midodrine is an oral medication that acts on alpha-1 adrenergic receptors in the peripheral vasculature and was initially approved by the FDA in 2010 for the treatment of symptomatic orthostatic hypotension despite uncertainty as to whether this translates to a proven clinical benefit in an ability to perform activities of daily living.^7^ Because of the hypotension associated with advanced heart failure and GDMT, midodrine is occasionally prescribed off-label to elevate mean blood pressure for symptomatic improvement and to allow for up-titration of GDMT. However, the prevalence of midodrine use in patients with congestive heart failure with reduced ejection fraction (HFrEF) is unknown. In very small studies from more than ten years ago, midodrine has been shown to improve heart failure symptoms and achieve higher doses of GDMT in patients with systolic CHF.^8,9^ However, the true effect of midodrine in patients with systolic CHF is unknown as it has not been studied in randomized trials or large observational studies. Currently, there are no American College of Cardiology (ACC), American Heart Association (AHA) or European Society of Cardiology recommendations for the use of midodrine in patients with CHF. We sought to investigate the use of midodrine in patients admitted to our tertiary care facility with systolic CHF to assess for GDMT prescription changes and clinical outcomes.

## Methods

This was a single-center retrospective cohort study evaluating the effect of midodrine on GDMT prescription patterns in patients admitted to our hospital with a primary diagnosis of decompensated HFrEF between January 2020 and December 2020. This study was approved by the University of South Florida Institutional Review Board (IRB Exempt, Study #001012). Inclusion criteria were adults aged 18 years or older, and hospitalization for a primary diagnosis of decompensated HFrEF with LVEF <50%. Exclusion criteria were LVEF ≥50%, end-stage renal disease (ESRD), history of or newly diagnosed orthostatic hypotension, death at index hospitalization, hospitalization not related to heart failure, and lack of follow-up by electronic health record (EHR). Orthostatic hypotension and ESRD were chosen as exclusion criteria to limit confounding as midodrine already has established uses for these conditions. For the midodrine cohort, an additional exclusion criterion was the administration of midodrine solely during the hospitalization and not continued upon discharge, to exclude transient midodrine use. TriNetX data of our institution was used initially to query the identification of patients with International Classification of Diseases-10 (ICD) codes based on the inclusion and exclusion criteria. TriNetX is a global health research network providing access to electronic medical record including diagnosis, procedure, laboratory data, medications. We retrieved the medical record number to perform individual chart review to confirm inclusion and exclusion criteria and clinical covariates. The midodrine cohort was then compared to consecutive patients over the same timeframe who presented with decompensated HFrEF who were not prescribed midodrine at any time between index admission and follow-up. Clinical data were collected by EHR review including demographics, all available cardiac imaging studies, and cardiac medications to include beta blockers, angiotensin-converting enzyme-inhibitors (ACE-i), angiotensin-receptor blockers (ARBs), ARNIs, mineralocorticoid receptor antagonists (MRAs), and sodium-glucose cotransporter-2 inhibitors (SGLT2i). The primary outcomes included the prevalence of GDMT at 6-month follow-up, prescription pattern, and dose adjustments. GDMT changes at 6 months were compared to the medication regimen upon discharge from index hospitalization, and were categorized as initiation, up-titration, discontinuation, decreased dose, or no change. We also evaluated for all-cause mortality, hospitalizations for heart failure, and the initiation of an advanced therapy with either a left ventricular assist device (LVAD) or orthotopic heart transplant (OHT) at 6 months.

### Statistical Analysis

Study data were collected and managed using REDCap electronic data capture tools hosted at the University of South Florida.^10^ The baseline characteristics and co-morbidities were presented as means. Comparison between the group was analyzed using the chi-square (χ^2^) test and Student’s *t-test* for categorical variables and continuous variables, respectively. Multivariate analysis was used to evaluate the association between midodrine use and all-cause mortality. The threshold for statistical significance was a 2-sided p<0.05. All analyses were performed using R Studio, version 4.0.4.

## Results

There were 668 admissions for systolic heart failure in the midodrine group of whom 114 met inclusion criteria, and 1712 admissions for systolic heart failure in the control group of whom 358 met inclusion criteria (Figure 1). The baseline characteristics of patients who were prescribed midodrine and those who were not are shown in Table 1. When compared to the control group, the patients who were prescribed midodrine had higher rates of baseline severe LV dysfunction, RV dysfunction, NYHA (New York Heart Association) class III and IV symptoms, AHA/ACC stage C or D heart failure, hypotension, and CKD (chronic kidney disease). They were also more likely to be on baseline inotropic support or have a pre-existing advanced therapy.

**Figure 1.**
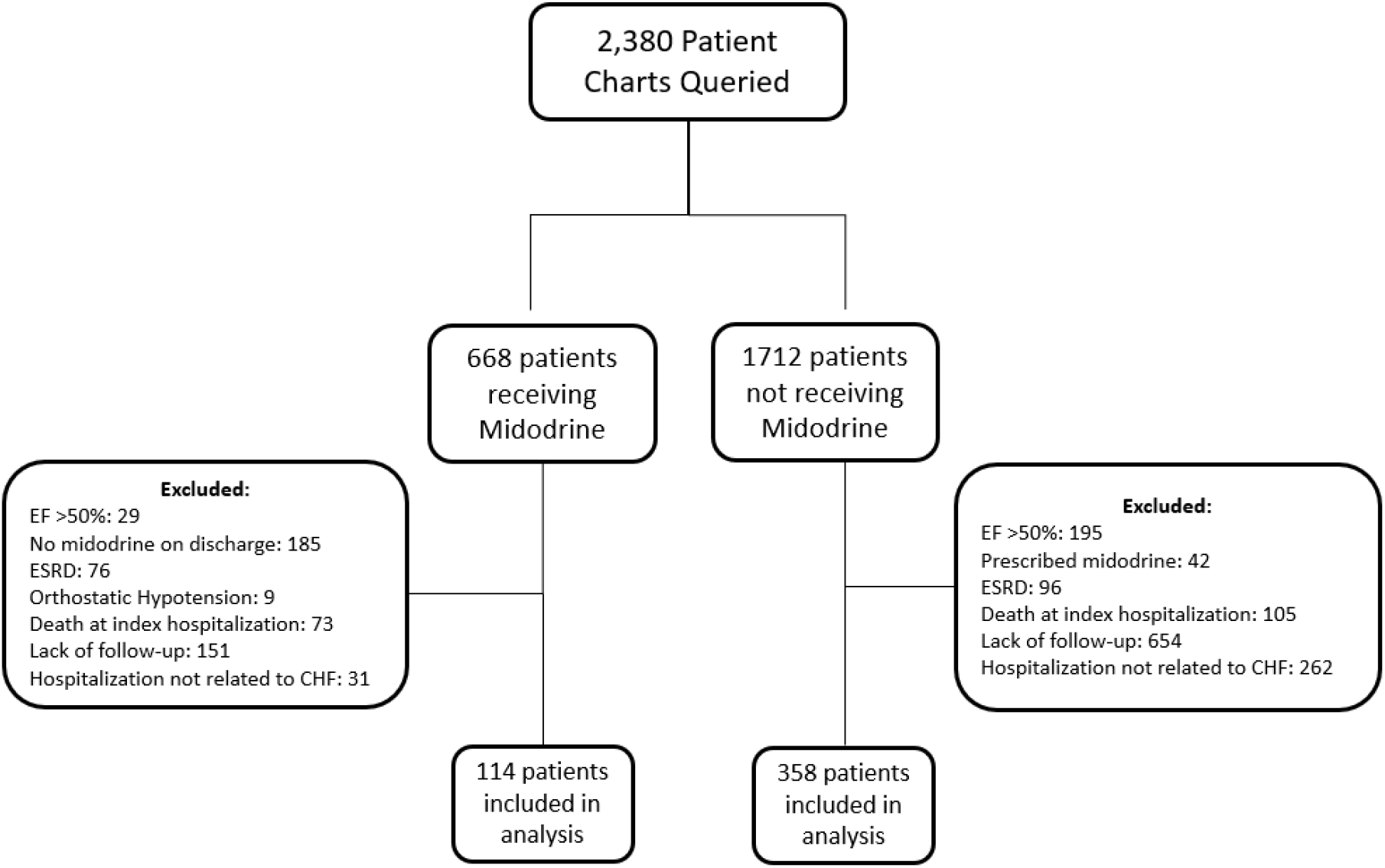
Inclusion and Exclusion of Patients in the Analysis. 2,380 patients were queried of which 668 were prescribed midodrine and 1712 were not. After individual chart review to confirm inclusion and exclusion criteria, 114 patients in the midodrine group and 358 patients in the non-midodrine group were included in the final analysis.

**Table 1.** Patient Demographic, Clinical Characteristics and Baseline Medical Therapy.

| Characteristic | Midodrine (n = 114) | No Midodrine (n = 358) | P-value |
| --- | --- | --- | --- |
| Age, mean± S.D, y | 61.6 ± 14.2 | 62.5 ± 14.8 | 0.559 |
| Male, No. (%) | 83 (72.8%) | 259 (72.3%) | 1 |
| Co-morbidities, No. (%) |  |  |  |
| CAD | 37 (32.5%) | 139 (38.8%) | 0.265 |
| Prior CABG/PCI | 43 (37.5%) | 133 (37.2%) | 1 |
| CKD | 52 (45.6%) | 102 (28.5%) | 0.001 |
| COPD | 20 (17.5%) | 59 (16.5%) | 0.904 |
| CVA | 24 (21.1%) | 47 (13.1%) | 0.056 |
| Diabetes | 50 (43.9%) | 151 (42.4%) | 0.836 |
| Hypertension | 55 (48.2%) | 272 (76.0%) | <0.001 |
| Obesity | 25 (21.9%) | 101 (28.2%) | 0.231 |
| Tobacco Use History | 26 (22.8%) | 61 (17.0%) | 0.213 |
| RV dysfunction | 57 (50.0%) | 132 (36.9%) | 0.017 |
| SBP, mean± S.D, mmHg | 99.3 ± 11.9 | 113.0 ± 17.4 | <0.001 |
| DBP, mean± S.D, mmHg | 61.5 ± 12.3 | 64.7 ± 14.8 | <0.022 |
| BMI, mean± S.D, kg/m<sup>2</sup> | 27.6 ± 7.3 | 29.9 ± 7.8 | 0.008 |
| BNP on arrival, mean± S.D , ng/L | 1280.8 ± 1398.2 | 1102.1 ± 1771.8 | 0.306 |
| NYHA Class, No. (%) |  |  | <0.001 |
| II | 5 (4.4%) | 98 (27.4%) |  |
| III | 76 (66.7%) | 235 (65.6%) |  |
| IV | 33 (28.9%) | 25 (7.0%) |  |
| LVEF group, No. (%) |  |  | <0.001 |
| 40-49% | 8 (7.0%) | 64 (18.1%) |  |
| 30-39% | 18 (15.8%) | 92 (26.0%) |  |
| 20-29% | 31 (27.2%) | 101 (28.5%) |  |
| <20% | 57 (50.0%) | 97 (27.4%) |  |
| Stage of Heart Failure |  |  | <0.001 |
| Stage B | 1 (0.9%) | 33 (9.2%) |  |
| Stage C | 57 (50.0%) | 291 (81.3%) |  |
| Stage D | 56 (49.1%) | 34 (9.5%) |  |
| Baseline Inotropic Support |  |  | <0.001 |
| Prior to Admission | 31 (27.4%) | 15 (4.2%) |  |
| New use | 26 (23.0%) | 23 (6.4%) |  |
| No use | 56 (49.6%) | 320 (89.4%) |  |
| Diuretic Use | 91 (79.8%) | 253 (70.6%) | 0.027 |
| New Diagnosis of HF During Index Admission | 8 (7.1%) | 63 (18.8%) | 0.005 |
| HF hospitalizations within last year |  |  | 0.015 |
| 0 | 54 (47.8%) | 212 (60.1%) |  |
| 1 | 23 (20.4%) | 76 (21.5%) |  |
| 2 | 20 (17.7%) | 30 (8.5%) |  |
| >2 | 16 (14.2%) | 35 (9.9%) |  |

Values are presented as number (Percentage) or mean +/- standard deviation based on type of data.
**Abbreviations:** coronary artery disease (CAD), coronary artery bypass graft (CABG), chronic kidney disease (CKD), chronic obstructive pulmonary disease (COPD), cerebrovascular accident (CVA), diastolic blood pressure (DBP), heart failure (HF), right ventricle (RV), body mass index (BMI), B-type natriuretic peptide (BNP), New York Heart Association (NYHA), left ventricular ejection fraction (LVEF), systolic blood pressure (SBP).

Baseline and follow-up GDMT prescription rates are shown in Table 2. Upon discharge from index hospitalization, the midodrine group, as compared to the control group had lower prescription rate of beta blockers (74.6% vs 89.4%) and ACE-i/ARBs (13.2% vs 32.4%), and similar ARNIs (37.7% vs 37.7%), MRAs (47.4% vs 44.7%) and SGLT2-is (30.7% vs 26.5%). At 6-month follow-up, patients who were prescribed midodrine were more likely to have initiation or up-titration of beta blockers (24.6% vs 15.4%; p=0.035), ACE/ARB/ARNIs (34.2% vs 24.0%; p=0.043) and SGLT2i (19.3% vs 10.6%; p=0.024) compared to the non-midodrine cohort, with a similar pattern for MRA prescriptions (17.5% vs 11.5%; p=0.126) which did not meet statistical significance.

**Table 2.**
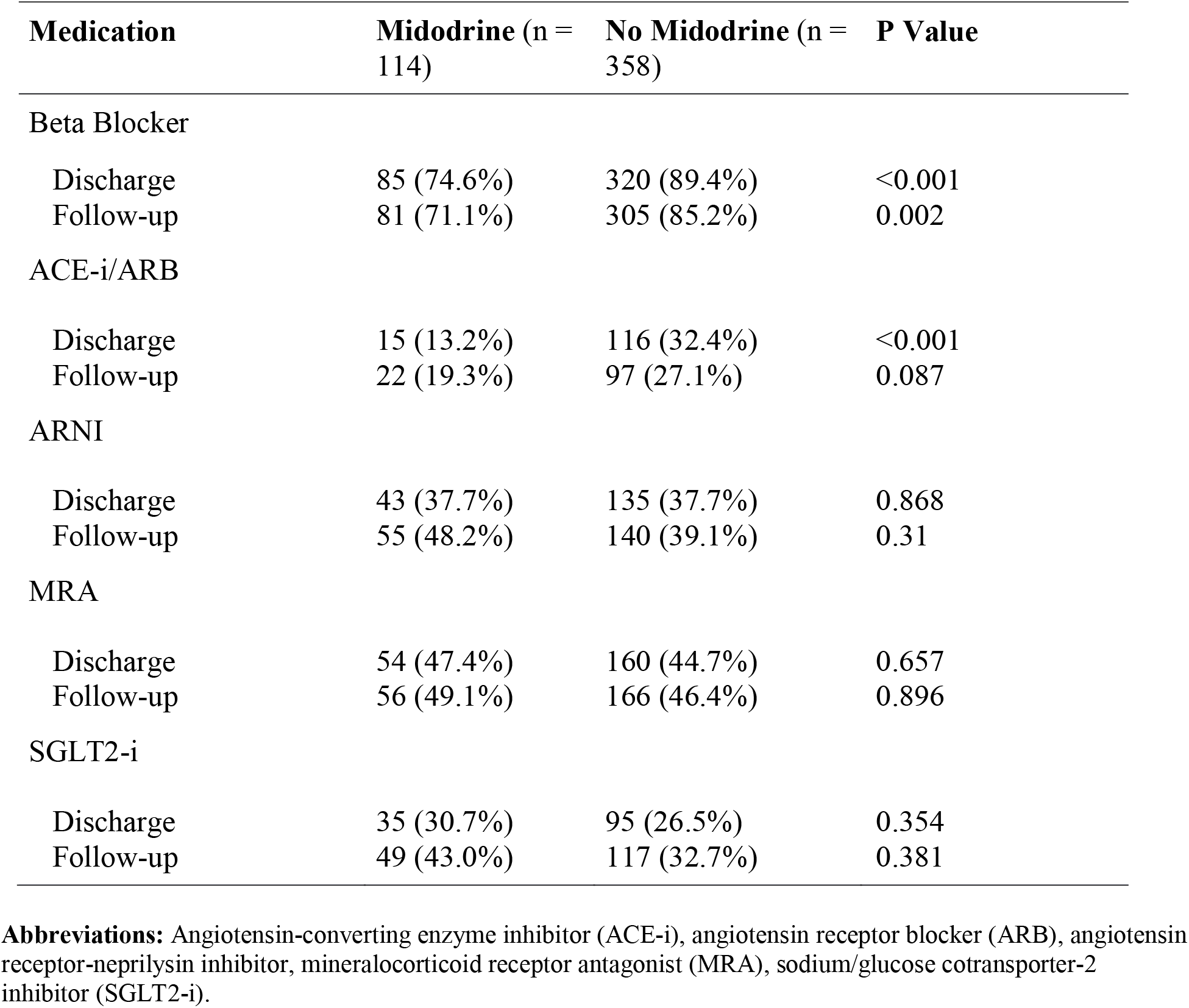
Guideline-Directed Medical Therapy at Discharge and 6-month Follow-Up.

| **Medication** | **Midodrine (n = 114)** | **No Midodrine (n = 358)** | **P Value** |
| --- | --- | --- | --- |
| **Beta Blocker** |  |  |  |
| Discharge | 85 (74.6%) | 320 (89.4%) | <0.001 |
| Follow-up | 81 (71.1%) | 305 (85.2%) | 0.002 |
| **ACE-i/ARB** |  |  |  |
| Discharge | 15 (13.2%) | 116 (32.4%) | <0.001 |
| Follow-up | 22 (19.3%) | 97 (27.1%) | 0.087 |
| **ARNI** |  |  |  |
| Discharge | 43 (37.7%) | 135 (37.7%) | 0.868 |
| Follow-up | 55 (48.2%) | 140 (39.1%) | 0.31 |
| **MRA** |  |  |  |
| Discharge | 54 (47.4%) | 160 (44.7%) | 0.657 |
| Follow-up | 56 (49.1%) | 166 (46.4%) | 0.896 |
| **SGLT2-i** |  |  |  |
| Discharge | 35 (30.7%) | 95 (26.5%) | 0.354 |
| Follow-up | 49 (43.0%) | 117 (32.7%) | 0.381 |

**Abbreviations:** Angiotensin-converting enzyme inhibitor (ACE-i), angiotensin receptor blocker (ARB), angiotensin receptor-neprilysin inhibitor, mineralocorticoid receptor antagonist (MRA), sodium/glucose cotransporter-2 inhibitor (SGLT2-i).

**Table 3.** Change in Guideline-Directed Medical Therapy At 6-Month Follow-Up.

| **Medication** | **Midodrine**<br>**(n = 114)** | **No Midodrine**<br>**(n = 358)** | **P-value** |
| --- | --- | --- | --- |
| Beta Blocker |  |  | 0.035 |
| Initiate/Uptitrate | 28 (24.6%) | 55 (15.4%) |  |
| Discontinue/Decrease/No change | 86 (75.4%) | 303 (84.6%) |  |
| ACE-i/ARB/ARNI |  |  | 0.043 |
| Initiate/Uptitrate | 39 (34.2%) | 86 (24.0%) |  |
| Discontinue/Decrease/No change | 75 (65.8%) | 272 (76.0%) |  |
| MRA |  |  | 0.126 |
| Initiate/Uptitrate | 20 (17.5%) | 41 (11.5%) |  |
| Discontinue/Decrease/No change | 94 (82.5%) | 317 (88.5%) |  |
| SGLT2-i |  |  | 0.024 |
| Initiate/Uptitrate | 22 (19.3%) | 38 (10.6%) |  |
| Discontinue/Decrease/No change | 92 (80.7%) | 320 (89.4%) |  |

**Abbreviations:** Angiotensin-converting enzyme inhibitor (ACE-i), angiotensin receptor blocker (ARB), angiotensin receptor-neprilysin inhibitor, mineralocorticoid receptor antagonist (MRA), sodium/glucose cotransporter-2 inhibitor (SGLT2-i).

**Table 4.** Clinical Outcomes at 6-Months.

| Outcome | Midodrine (n=114) | No Midodrine (n=358) | P-value |
| --- | --- | --- | --- |
| Survival | 97 (85.1%) | 345 (96.4%) | < 0.001 |
| Advanced Therapies |  |  | < 0.001 |
| Heart Transplant | 9 (7.9%) | 10 (2.8%) |  |
| LVAD | 7 (6.1%) | 13 (3.6%) |  |
| Inotrope Use | 36 (31.6%) | 24 (6.7%) |  |
| HF Readmission |  |  | 0.006 |
| 0-1 | 69 (60.5%) | 267 (74.6%) |  |
| 2+ | 45 (39.5%) | 91 (25.4%) |  |

Abbreviations: LVAD: left ventricular assist device; HF: heart failure

All-cause mortality at 6-months was significantly higher in the midodrine group as compared to the group without midodrine (Odds Ratio [OR] 4.65, 95% CI 2.18-9.91, p<0.001), however when adjusting for relevant clinical factors including baseline blood pressure, chronic kidney disease, NYHA classification and baseline inotrope status, the overall mortality was not statistically different (OR 2.26, 95% CI 0.90-5.68, p=0.083). Frequent re-hospitalization for heart failure was more common in the midodrine group compared to control (39.5% vs 25.4%; p=0.006). Patients who were prescribed midodrine were significantly more likely to receive an LVAD or OHT as compared to those not prescribed midodrine (14% vs 6.4%; p<0.001).

## Discussion

The ability to administer GDMT to patients with systolic heart failure is frequently opposed by hypotension, and previous studies have demonstrated clinician hesitance to adequately up-titrate pharmacotherapy for this reason.^11^ We found that the use of midodrine is associated with the ability to initiate or up-titrate GDMT among the systolic heart failure population, particularly with beta blockers and RAAS-inhibitors. These findings are highly relevant as recent randomized trials have documented significant reductions in death and heart failure hospitalizations when GDMT is rapidly uptitrated.^12^ Surprisingly, both groups had a small decrease in overall beta blocker usage at 6 months, which indicates that there were slightly more discontinuations than initiations in both groups. The reasons for beta blocker discontinuation in the midodrine group, as compared with the non-midodrine group, included advanced therapy initiation (68% vs. 28%) and medication intolerance or patient refusal (32% vs. 72%).

One particularly striking finding, however, was the frequency with which midodrine has been applied to these patients at all—despite lack of convincing evidentiary support. To wit, multiple studies have previously suggested harm of midodrine in critically ill patients.^13,14^ Almost 25% of included patients were prescribed midodrine; this important finding underscores the need for future prospective research into the matter.

We also identified many notable baseline characteristics differences between the two cohorts. The baseline features of the typical patient likely to receive midodrine include more severe LV dysfunction, more frequent RV dysfunction, more severe symptoms as assessed by NYHA classification and were more likely to require loop diuretics for maintenance therapy. The patients in the midodrine cohort had overall worse clinical prognosis, including worse survival (although not statistically significant) and rehospitalization for heart failure. These findings should be interpreted with caution as they may simply reflect the higher baseline rates of LV and RV dysfunction, which limit the ability to interpret the effect of midodrine. However there is biologic plausibility for worse outcomes on midodrine as, for the failing heart, the counterproductive increased systemic vascular resistance may outmatch the benefits of improved pharmacotherapy. The patients who were prescribed midodrine were also more likely to undergo some form of advanced cardiac therapy (LVAD, OHT) at 6-months. The reasons for this are commensurate with more advanced stages of disease in the midodrine cohort. As midodrine is occasionally prescribed as a reactive clinical maneuver for treating symptomatic hypotension with the secondary benefit of allowing GDMT titration, the use of midodrine in this population likely reflects the branding of a subgroup within the low BP population that requires earlier and aggressive attention. This is important because intermittent or marginal hypotension in the advanced heart failure patient might go overlooked, whereas a midodrine prescription may more quickly signify to the provider to consider an advanced therapy referral. Current ACC expert consensus includes the acronym “I-NEED-HELP” as a tool to quickly determine which patients may benefit from referral to a heart failure specialist.^5^ We suspect that the prescription of midodrine may warrant similar early referrals for advanced cardiac therapies.

Our study has numerous important limitations. As with all retrospective reviews, we make no causative claims about midodrine and GDMT prescription patterns or mortality—only its association, which may have confounding variables not accounted for in this study. While it is possible that the superior GDMT titration in the midodrine arm could be explained by more frequent medical contact, all patients had to have physician follow-up to be included in the study and thus reflects real-world GDMT adjustments. In addition, we are only able to track survival and rehospitalization if the information is available to our institution or those institutions that share EHR data with our network. Second, we excluded patients with a lack of follow-up or confirmation of death which has the potential to bias results; however, this occurred in both cohorts at similar rates, and we expect the effect to be minimal. Third, being a single-center experience, our prescription patterns for midodrine use may differ significantly from other centers; however, our institution has a high volume of cardiac transplantations and mechanical circulatory support implantations, and therefore our findings may be highly relevant for similar programs. Despite the limitations, we have notable strength in our study. Our study is novel as there is no large-scale study describing the effect of midodrine on prescription patterns for GDMT of heart failure. We clearly see increased utilization of GDMT in the midodrine group.

Future efforts in evaluating the effects of midodrine in advanced heart failure patients should be performed in prospective clinical trials.

## Conclusion

In our study, we observed that patients admitted with systolic heart failure who were prescribed midodrine achieved better initiation and up-titration of GDMT at 6 months compared to those who were not prescribed midodrine. The cohort of patients prescribed midodrine represents a population with more advanced heart failure and worse clinical outcomes, reflecting a subgroup that warrants consideration for earlier and aggressive intervention.

## Data Availability

All data produced in the present study are available upon reasonable request to the authors.

## Abbreviations

ACC: (American College of Cardiology)
AHA: (American Heart Association)
NYHA: (New York Heart Association)
(CHF): Congestive heart failure
(GDMT): Guideline directed medical therapy
(HFrEF): Heart failure with reduced ejection fraction
LVAD: (left ventricular assist device)
(LVEF): Left ventricular ejection fraction
OHT: (orthotopic heart transplant)

## Acknowledgements

A portion of Figure 1 was created with BioRender.com. TriNetX was utilized to query for patient charts.

